# The effect of dysautonomia on motor, behavioral and cognitive fluctuations in Parkinson’s disease

**DOI:** 10.1101/2024.08.26.24312589

**Authors:** Abhimanyu Mahajan, Christopher B Morrow, Joseph Seemiller, Kelly A Mills, Gregory M Pontone

## Abstract

**Background:** Motor and non-motor fluctuations adversely impact quality of life in Parkinson’s disease (PD). Dysautonomia, a feature frequently associated with PD and a possible adverse effect of dopaminergic therapy, may be comorbid with fluctuations.

**Objectives:** We sought to evaluate the effect of dysautonomia on motor and non-motor fluctuations in PD

**Methods:** Two hundred subjects with PD were evaluated in both “on” and “off” dopamine states to assess changes in symptoms related to dopaminergic fluctuations. Multivariable logistic regression was performed to assess the association of dysautonomia with motor, cognitive, and psychiatric worsening from ON to OFF states with adjustment for disease duration, levodopa equivalent daily dosage (LEDD), and dopamine agonist LEDD.

**Results:** Subjects with dysautonomia had greater odds of clinically meaningful change in motor features (OR 3.0), cognition (OR 3.4) and anxiety (OR 4.3) compared to those without dysautonomia.

**Conclusions:** Dysautonomia may be a contributory mechanism behind fluctuations in PD. The exact nature of this relationship deserves further evaluation.

## Introduction

Dysautonomia, defined as impairment of the autonomic nervous system, is commonly seen in patients with PD. It may also be seen in undiagnosed patients at high-risk for developing motor manifestations of PD.^1^ A high prevalence of different aspects of dysautonomia has been reported in patients with PD: 20-60% with urinary or sexual dysautonomia, 10-15% with thermoregulatory dysfunction, 19-50% with gastrointestinal dysfunction, and 10-20% with pupillomotor disorders.^2-5^ Orthostatic hypotension and supine hypertension may be seen in 20-40% of PD patients with a profound impact on falls, quality of life and a greater need for medical care.^5, 6^ Even if asymptomatic, it may affect overall function and quality of life.^7^

A robust response to dopaminergic therapy is a hallmark of idiopathic Parkinson’s disease (PD). However, motor fluctuations in response to treatment may be seen in approximately 40-50% of PD patients 5 years into the disease, with disease duration and treatment dose as predictors.^8^ Motor fluctuations typically present as an initial end-of-dose worsening of motor features and may progress to unpredictable fluctuations with a profound impact on quality of life.^9^ In addition, PD patients with motor fluctuations may report non-motor fluctuations in anxiety (35.4%), depressive symptoms (34.9%) and feelings of panic (37.1%).^10-12^ While initially believed to be a consequence of motor fluctuations, noted discordance between severity of fluctuations in motor features and mood led to the acceptance of at least, partial independence between motor and non-motor fluctuations.^13, 14^ Such non-motor fluctuations have an additional, substantive impact on quality of life in PD and might not resolve with treatment of motor fluctuations.^15^

Dysautonomia is known to worsen with dopaminergic therapy and may be comorbid with fluctuations.^16^ Whether dysautonomia is associated with, or even potentiates, motor and non-motor fluctuations is poorly understood.

Through this effort, we sought to evaluate whether the presence of dysautonomia in persons with PD is associated with the degree to which dopamine replacement affects motor and non-motor function, with specific interest on its impact on changes in mood. Such a relationship, if demonstrated, may suggest aggressive targeting of dysautonomia may impact the experience of motor or non-motor fluctuations.

## Methods

### Participants

The study protocol^17^ included adults 21 years of age or older, diagnosed with idiopathic Parkinson’s disease based on MDS clinical diagnostic criteria^18^. Those in the first year of illness and those diagnosed with dementia were excluded to avoid enrolling individuals with Lewy body dementia. No cognitive cut-off was used to avoid excluding patients with advanced PD who were still able to function independently and did not meet criteria for dementia. Recruitment sites included the Morris K. Udall Parkinson’s Disease Research Center at the Johns Hopkins University School of Medicine, the Johns Hopkins Parkinson’s Disease Research Center, and three outpatient movement disorder neurology clinics. A total of 200 participants completed the study and were included in the analysis.

### Measures

Dysautonomia was defined using the Scales for Outcomes in Parkinson’s Disease – Autonomic Dysfunction (SCOPA-AUT), a well validated scale for measuring symptoms of autonomic dysfunction in PD over the previous month.^19^ The established cut-off score of 13.1 was used to define those with dysautonomia.^4^ Mood symptoms were measured using the Hamilton Anxiety Rating Scale (HAM-A) and the Hamilton Depression Rating Scale (HAM-D), which are well-established measures of anxiety and depressive symptoms commonly used and validated in populations with PD.^20^ The HAM-A is a 17-item rating scale that measures the presence and intensity of different anxiety symptom domains. Motor functioning was assessed using the Movement Disorders Society-Unified Parkinson’s Disease Rating Scale Part III (MDS-UPDRS III).^21^ Cognitive measures were administered and scored according to standard instructions by a trained research assistant. The Symbol Digit Modalities Test (SDMT) and STROOP test were administered in an oral format to avoid motor confounds associated with the written format. Total and dopamine agonist-associated levodopa equivalent daily dosage (LEDD) was calculated for each participant.^11^

### Procedures

Participants were evaluated in both “on” and “off” dopamine states to assess changes in symptoms related to dopaminergic fluctuations. Participants were first evaluated after stopping all dopaminergic medications for a minimum of 12 hours to establish an “off” medication state baseline. Participants then took their typical morning dopaminergic medications and were re-evaluated once they reported being in their best “on” medication state. The change in mood symptoms between “on” and “off” dopamine states was calculated. Assessments were conducted by Movement Disorder Society (MDS) trained clinicians in an outpatient research setting. The Johns Hopkins University Institutional Review Board approved the study protocol, and all participants provided written informed consent before participation.

### Statistical Analysis

Differences in participant characteristics and clinical outcomes were compared using two-sample t-tests for continuous variables and Pearson χ2 tests for categorical variables. We performed multivariable logistic regression to assess the association of dysautonomia with motor, cognitive, and psychiatric worsening from ON to OFF states with adjustment for disease duration, levodopa equivalent daily dosage (LEDD), and dopamine agonist LEDD. Binary outcomes were defined based on established minimally clinically important differences (MCID) –a worsening of 4.63 on the MDS UPDRS III, 4 on the SDMT, 5.5 on thr STROOP test, and 4 on the HAM-D.^22-25^ There are no widely accepted MCID thresholds for the HAM-A, so we established a threshold of 7.5 which is 0.5 standard deviation beyond the mean change in our population and represents a meaningful increase in clinical anxiety. Alpha was set at 0.05. STATA SE 18 (StataCorp LP, College Station, TX) was used for all analyses.

## Results

### Demographic and Clinical Characteristics

Table 1 displays demographic, cognitive, motor, and psychiatric symptoms stratified by dysautonomia status for the 200 participants. Of the 200 participants, 106 exhibited symptoms of dysautonomia (SCOPA-AUT ≥ 13.1) constituting 53% of the study sample. Given the known relationship between dopamine replacement therapy and orthostatic hypotention, a sensitivity analysis excluding SCOPA-AUT items associated with orthostasis (questions 14-18) was conducted but did not materially alter the results. As such, symptoms related to orthostatic hypotension alone were not thought to be responsible for the relationship. SCOPA-AUT scores were calculated using all questions. There were no statistically significant differences in age, sex, or race in those with or without dysautonomia. There were no statistically significant cognitive differences between the groups. Duration of disease (8.2 versus 5.0, p<0.001), LEDD (877.6 versus 723.6, p<0.001), and dopamine-agonist LEDD (113.9 versus 61.3, p = 0.008) were all higher in the dysautonomia group.

**Table 1:** Demographic, Cognitive, Behavioral, and Motor Symptoms by Dysautonomia Status.

|  | **DYS-**<br>**(n = 94)** | **DYS+**<br>**(n = 106)** | **t-value**<br>**or $\chi^2$\*** | **p-value** |
| --- | --- | --- | --- | --- |
| **Demographics** | % | % |  |  |
| **Age** | 64.6 (7.9) | 65.7 (7.6) | -1 | 0.34 |
| **Sex (% male)** | 37.2 | 40.6 | 0.23 | 0.63 |
| **Race (% non-white)** | 5.3 | 9.4 | 1.22 | 0.27 |
| **Duration of disease** | 7.1 (4.8) | 10.8 (6.1) | -4.7 | <0.001 |
| **Levodopa equivalent dosage**<br>**(mg/day) [mean (SD)]** | 723.6<br>(450.3) | 877.6<br>(529.7) | -2.2 | 0.03 |
| **Agonist Levodopa equivalent**<br>**dosage (mg/day) [mean (SD)]** | 61.3<br>(107.7) | 113.9<br>(159.4) | -2.7 | 0.008 |
| **Cognition** |  |  |  |  |
| **MoCA score** | 27.1 (2.7) | 26.4 (3.1) | 1.5 | 0.13 |
| **SDMT score difference (post**<br>**vs. pre-treatment)** | -2.3 (5.9) | -0.9 (6.6) | -1.5 | 0.13 |
| **Motor Symptoms** |  |  |  |  |
| **MDS UPDRS III score difference**<br>**(post vs. pre-treatment)** | -12.2 (8.1) | -14.3 (7.6) | 1.9 | 0.05 |
| **Psychiatric Symptoms** |  |  |  |  |
| **HAM-A score difference (post**<br>**vs. pre-treatment)** | -4.0 (4.7) | -6.0 (4.8) | 2.9 | 0.004 |
| **HAM-D score difference (post vs. pre-treatment)** | -1.5 (2.6) | -2.3 (2.6) | 2.1 | 0.04 |

\*t-value for continuous variables, chi-square for binary variables. Abbreviations: MoCA: Montreal Cognitive Assessment, SDMT: Symbol Digit Modalities Test, Movement Disorders Society- Unified Parkinson's Disease Rating Scale (MDS-UPDRS) part III, HAM-A: Hamilton Anxiety Rating Scale, HAM-D: Hamilton Depression Rating Scale

The magnitude of improvement in motor (MDS UPDRS III change of -14.3 versus -12.2, p = 0.045) and anxiety symptoms (HAM-A change -6.0 versus -4.0, p<0.001) between on and off states was greater in the dysautonomia group relative to the non-dysautonomia group. There was no evidence of a difference in depressive symptom change between the groups. The odds of clinically meaningfully worsening in those with dysautonomia versus those without dysautonomia was 3.0 (95% CI 1.0 – 8.8) for motor symptoms, 3.4 (95% CI 1.5-7.9) for SDMT, and 4.3 (95% CI 1.9 – 9.9) for anxiety symptoms (Table 2, Supplementary data). Additional assessment of the relationship using linear regression models led to similar conclusions. An increase in SCOPA scores was associated with a significant difference in STROOP scores with medication. [-0.17 (0.07), p = 0.03) (Supplementary data: SupplementaryTable 1)

**Table 2:**
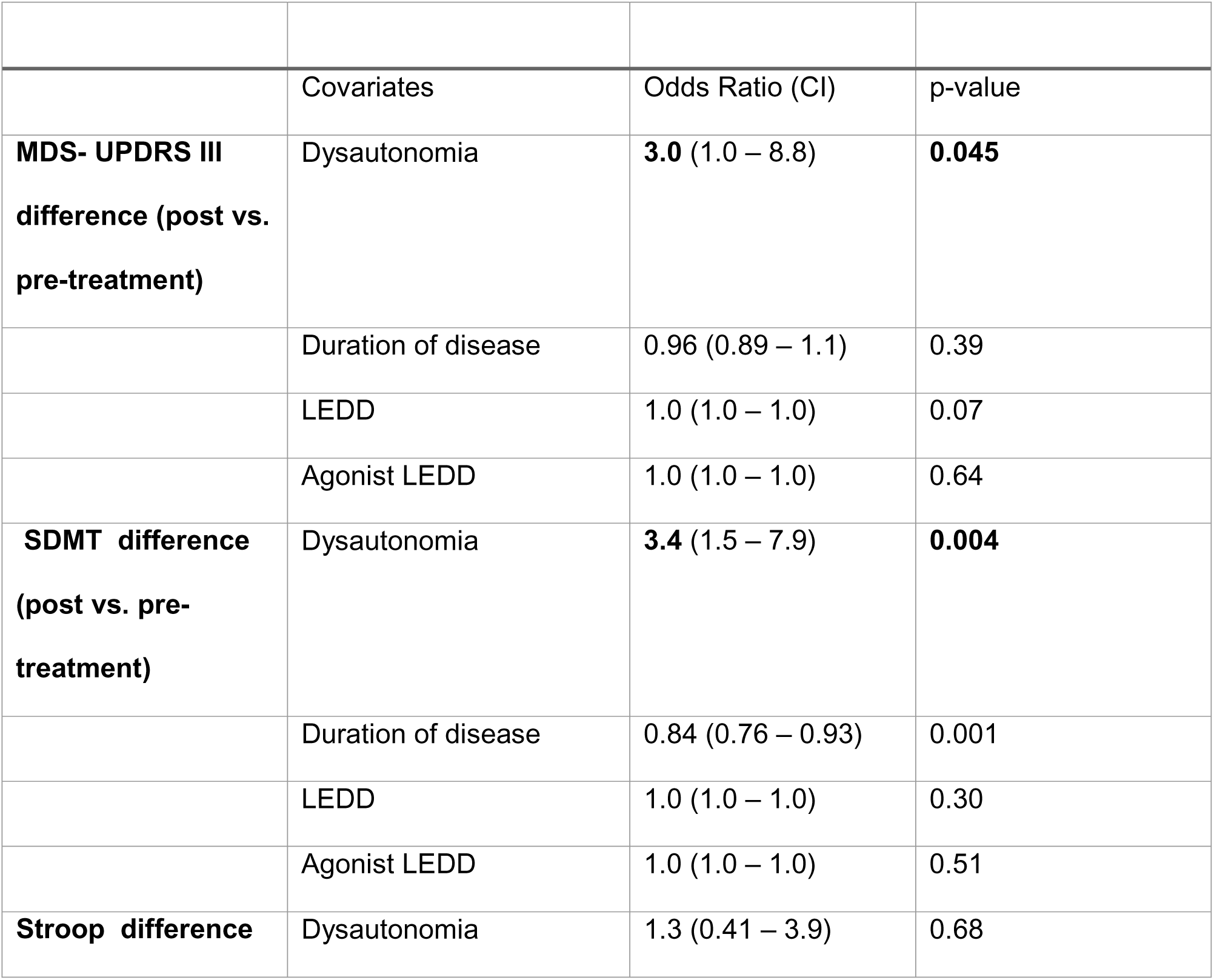
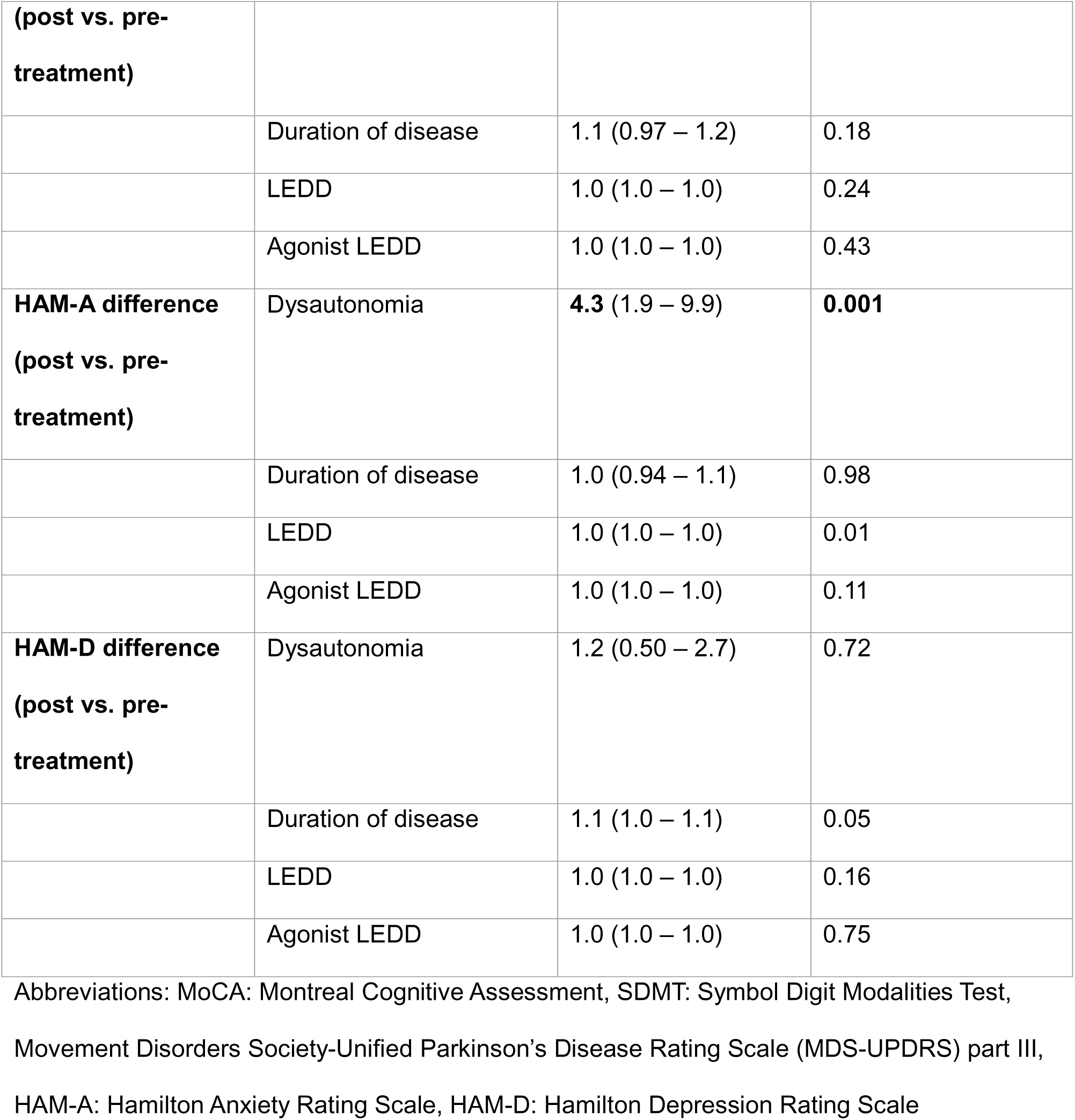
Odds of motor and non-motor fluctuations in Parkinson’s disease with dysautonomia.

## Discussion

Early reports of fluctuations with dopaminergic therapy included the co-prevalence of dysautonomia and behavioral/ cognitive features with descriptions of flushing, sweating, fear and confusion, along with mood swings.^14, 26^ In our sample of 200 patients with systematically collected data before and after dopaminergic treatment, the presence of dysautonomia over the last month was associated with greater odds of clinically-relevant change in anxiety, cognition (SDMT: psychomotor speed, attention, visual scanning and tracking, and working memory)^27^ and motor features.

Motor fluctuations are thought to represent the combined effect of pulsatile dopaminergic stimulation and disease progression, leading to a reduction in dopamine synthesis and presynaptic dopaminergic terminal storage.^14, 28, 29^ Additionally, post-synaptic changes, as a consequence of disease and treatment, likely contribute.^14^ Despite shared risk factors of disease duration, treatment dose and duration, and PD progression, non-motor fluctuations are not entirely explained by the kinetics of dopamine repletion as it relates to motor function.^13, 14^ Thus, other mechanisms may affect the relationship between dopamine replacement and non-motor symptom changes. Our data may suggest that, accounting for disease duration and dopamine replacement dose as surrogates of overall disease severity, the presence of dysautonomia may predispose individuals to experience greater differences in anxiety with dopamine repletion.

In support of this notion, several other studies suggest a causal role of dysautonomia in the experience of anxiety in PD. Dysautonomia has been reported to be an independent predictor of anxiety in PD.^30^ Greater dysautonomia, especially gastrointestinal and thermoregulatory symptoms, has been associated with greater mood related symptom burden longitudinally in early PD.^31^ A technique called heart rate variability biofeedback, involving self-regulation of a physiological dysregulated vagal nerve function, especially when combined with cognitive behavioral therapy has been reported to improve anxiety.^32^ While a possible explanation is that manifestations of dysautonomia (tachycardia, reduced heart rate variability) overlap with self-reported anxiety, ^30, 33^ the role of the autonomic nervous system in anxiety is well established with substantial reductions in heart rate variability (poor vagal control) noted across psychiatric disorders in medication-free individuals.^34^ A recent study offered preliminary evidence of cardiac sympathetic denervation explaining ∼20% of pure anxiety variance in PD, independent of sex, dopaminergic impairment, and anxiolytic treatments.^35^ As such, dysautonomia may be postulated as a cause of poor response to anti-anxiety medications in some individuals.^36^

A similar relationship has been proposed with cognition.^37^ A review of relevant studies with direct and indirect evidence reported a relationship between dysautonomia and cognitive impairment. ^38^ Dysautonomia, manifesting as early orthostatic hypotension, is shown to predict greater cognitive dysfunction.^39^ PD patients with orthostatic hypotension decline faster in executive function compared to those with normal blood pressures.^40^ The relationship between OH and cognitive decline may be explained by widespread peripheral and central noradrenergic loss.^37^ Noradrenaline modulates cognition, specifically attention and working memory.^41^ Alpha-synuclein burden in Locus Coeruleus, a key source of noradrenaline, precedes and is noted to be of a greater magnitude than substantia nigra.^42^ Dysautonomia-mediated cerebral hypoperfusion has also been postulated as a contributing mechanism.^38^ Fluctuations in cognition have been associated with blood pressure fluctuation with clinical presentation^43^ and results of neuropsychological testing^44^ in PD. Improvement in orthostatic hypotension has been postulated as a mechanism of clinical benefit seen with use of Rivastigmine in cognitively impaired PD patients.^45^ Our study results, suggesting a contribution of dysautonomia to clinically-relevant fluctuations in anxiety in a robust sample of subjects with PD, are consistent with this literature. While linear regression modeling showed a statistically significant effect of dysautonomia on STROOP and SDMT scores, logistic regression modeling (with an additional focus on clinically-meaningful significance) only showed a relationship between dysautonomia and SDMT. An effect of dopaminergic treatment on SDMT performance has been demonstrated using the same data set and is thought to be potentially related to excess dopaminergic stimulation along circuits innervating the pre-frontal cortex.^46^ These conflicting results are consistent with literature with previous studies showing no effect of dopaminergic medications on STROOP test.^47-49^ Both SDMT and STROOP are tests that measure processing speed, workingmemory, attention, and impulse suppression. However, subtle differences remain including the assessment of speed of visual search, word reading and color naming. The effect of dysautonomia on cognitive fluctuations needs additional, nuanced assessment.

Our study sample with a mean age of 65.2 years (SD. 7.7 years), mean disease duration of 9.1 years (SD. 5.8 years) and LEDD of 805.2 mg (SD. 498.7 mg) is a reasonable representation of PD in the clinic. Our model accounted for dopamine-agonist dose which may disproportionately impact studied outcomes. The use of clinically relevant change in outcomes, representative of clinical impact, is a strength of our analysis. Our study has several limitations. On and Off states were defined based on withdrawal of dopaminergic medication. A rationale of this approach is its use for determination of candidacy for deep brain stimulation in PD.^50^ However, this manufactured off-state may differ from spontaneous or unpredictable fluctuations as any uncertainty associated with such fluctuations may be associated with greater anxiety. Moreover, our battery to assess non-motor features was limited given sample size constraints. The use of HAM-A and HAM-D to assess changes in the on- and off-states in PD has not been validated. Similarly, SDMT and STROOP scales have not been validated for this purpose. Genetic status of included subjects was not available. As such, the differential effect of genetic mutations on dysautonomia, and motor and non-motor fluctuations and their relationship cannot be accounted for. Regardless, the study design allows for a robust assessment of the impact of dysautonomia, a common manifestation in idiopathic PD, on dopamine-replacement association changes in motor and non-motor manifestations. In addition to offering a potential insight into a mechanism behind the reported relationship, it offers a potential therapeutic avenue for management of motor and non-motor fluctuations by addressing dysautonomia to target non-motor fluctuations in mood. This direction must be explored further in clinical studies.

## Data Availability

All data produced in the present study are available upon reasonable request to the authors

## Acknowledgement

We would like to thank the subjects for participating in this study.

## Ethical Compliance Statement

The Johns Hopkins University Institutional Review Board approved the study protocol. The research team obtained written consent from all participants prior to their participation. We confirm that we have read the journal’s position on issues involved in ethical publication and affirm that this work is consistent with those guidelines.

## Author Contributions

1. Research project: A. Conception, B. Organization, C. Execution;
2. Statistical Analysis: A. Design, B. Execution, C. Review and Critique;
3. Manuscript: A. Writing of the first draft, B. Review and Critique.

AM: 1A, 1B, 1C, 2A, 2C, 3A, 3B

CBM: 1B, 1C, 2A, 2B, 2C, 3B

JS: 2C, 3B

KAM: 2A, 2C, 3B

GMP: 1B, 1C, 2C, 3B

## Disclosures

### Financial disclosures from the last 12 months

The authors declare that there are no additional relevant disclosures to report

## References

1. Liepelt-Scarfone I, Pilotto A, Müller K, et al. Autonomic dysfunction in subjects at high risk for Parkinson’s disease. J Neurol. 2015 Dec;262(12):2643–52.

2. Chaudhuri KR, Prieto-Jurcynska C, Naidu Y, et al. The nondeclaration of nonmotor symptoms of Parkinson’s disease to health care professionals: an international study using the nonmotor symptoms questionnaire. Movement disorders : official journal of the Movement Disorder Society. 2010 Apr 30;25(6):704–9.

3. Wüllner U, Schmitz-Hübsch T, Antony G, et al. Autonomic dysfunction in 3414 Parkinson’s disease patients enrolled in the German Network on Parkinson’s disease (KNP e.V.): the effect of ageing. Eur J Neurol. 2007 Dec;14(12):1405–8.

4. Arnao V, Cinturino A, Valentino F, et al. In patient’s with Parkinson disease, autonomic symptoms are frequent and associated with other non-motor symptoms. Clin Auton Res. 2015 Oct;25(5):301–7.

5. Merola A, Romagnolo A, Rosso M, et al. Autonomic dysfunction in Parkinson’s disease: A prospective cohort study. Movement disorders : official journal of the Movement Disorder Society. 2018 Mar;33(3):391–7.

6. Espay AJ, LeWitt PA, Hauser RA, Merola A, Masellis M, Lang AE. Neurogenic orthostatic hypotension and supine hypertension in Parkinson’s disease and related synucleinopathies: prioritisation of treatment targets. Lancet Neurol. 2016 Aug;15(9):954–66.

7. Merola A, Romagnolo A, Rosso M, et al. Orthostatic hypotension in Parkinson’s disease: Does it matter if asymptomatic? Parkinsonism Relat Disord. 2016 Dec;33:65–71.

8. Schrag A, Quinn N. Dyskinesias and motor fluctuations in Parkinson’s disease. A community-based study. Brain. 2000 Nov;123 ( Pt 11):2297–305.

9. Hechtner MC, Vogt T, Zöllner Y, et al. Quality of life in Parkinson’s disease patients with motor fluctuations and dyskinesias in five European countries. Parkinsonism Relat Disord. 2014 Sep;20(9):969–74.

10. van der Velden RMJ, Broen MPG, Kuijf ML, Leentjens AFG. Frequency of mood and anxiety fluctuations in Parkinson’s disease patients with motor fluctuations: A systematic review. Movement disorders : official journal of the Movement Disorder Society. 2018 Oct;33(10):1521–7.

11. Pontone GM, Perepezko KM, Hinkle JT, et al. ’Anxious fluctuators’ a subgroup of Parkinson’s disease with high anxiety and problematic on-off fluctuations. Parkinsonism Relat Disord. 2022 Dec;105:62–8.

12. Dissanayaka NN, Forbes EJ, Perepezko K, et al. Phenomenology of Atypical Anxiety Disorders in Parkinson’s Disease: A Systematic Review. Am J Geriatr Psychiatry. 2022 Sep;30(9):1026–50.

13. Maricle RA, Nutt JG, Valentine RJ, Carter JH. Dose-response relationship of levodopa with mood and anxiety in fluctuating Parkinson’s disease: a double-blind, placebo-controlled study. Neurology. 1995 Sep;45(9):1757–60.

14. Martínez-Fernández R, Schmitt E, Martinez-Martin P, Krack P. The hidden sister of motor fluctuations in Parkinson’s disease: A review on nonmotor fluctuations. Movement disorders : official journal of the Movement Disorder Society. 2016 Aug;31(8):1080–94.

15. Kakimoto A, Kawazoe M, Kurihara K, Mishima T, Tsuboi Y. Impact of non-motor fluctuations on QOL in patients with Parkinson’s disease. Front Neurol. 2023;14:1149615.

16. Witjas T, Kaphan E, Azulay JP, et al. Nonmotor fluctuations in Parkinson’s disease: frequent and disabling. Neurology. 2002 Aug 13;59(3):408–13.

17. NIHreporter. Anxiety in Parkinson’s: Use of quantitative methods to guide rational treatment. 2014 [June 6, 2024]; Available from: https://reporter.nih.gov/search/AyzaeM4JmESgTldq1ht9gA/project-details/8634954.

18. Postuma RB, Berg D, Stern M, et al. MDS clinical diagnostic criteria for Parkinson’s disease. Movement disorders : official journal of the Movement Disorder Society. 2015 Oct;30(12):1591–601.

19. Visser M, Marinus J, Stiggelbout AM, Van Hilten JJ. Assessment of autonomic dysfunction in Parkinson’s disease: the SCOPA-AUT. Mov Disord. 2004 Nov;19(11):1306–12.

20. Leentjens AF, Dujardin K, Marsh L, Richard IH, Starkstein SE, Martinez-Martin P. Anxiety rating scales in Parkinson’s disease: a validation study of the Hamilton anxiety rating scale, the Beck anxiety inventory, and the hospital anxiety and depression scale. Movement disorders : official journal of the Movement Disorder Society. 2011 Feb 15;26(3):407–15.

21. Goetz CG, Tilley BC, Shaftman SR, et al. Movement Disorder Society-sponsored revision of the Unified Parkinson’s Disease Rating Scale (MDS-UPDRS): scale presentation and clinimetric testing results. Mov Disord. 2008 Nov 15;23(15):2129–70.

22. Horvath K, Aschermann Z, Acs P, et al. Minimal clinically important difference on the Motor Examination part of MDS-UPDRS. Parkinsonism Relat Disord. 2015 Dec;21(12):1421–6.

23. Rush AJ, South C, Jain S, et al. Clinically Significant Changes in the 17- and 6-Item Hamilton Rating Scales for Depression: A STAR*D Report. Neuropsychiatr Dis Treat. 2021;17:2333–45.

24. Benedict RH, DeLuca J, Phillips G, et al. Validity of the Symbol Digit Modalities Test as a cognition performance outcome measure for multiple sclerosis. Mult Scler. 2017 Apr;23(5):721–33.

25. Borland E, Edgar C, Stomrud E, Cullen N, Hansson O, Palmqvist S. Clinically Relevant Changes for Cognitive Outcomes in Preclinical and Prodromal Cognitive Stages: Implications for Clinical Alzheimer Trials. Neurology. 2022 Sep 13;99(11):e1142–e53.

26. Marsden CD, Parkes JD. "On-off" effects in patients with Parkinson’s disease on chronic levodopa therapy. Lancet. 1976 Feb 7;1(7954):292-6.

27. Ryan J, Woods RL, Britt CJ, et al. Normative Data for the Symbol Digit Modalities Test in Older White Australians and Americans, African-Americans, and Hispanic/Latinos. J Alzheimers Dis Rep. 2020 Aug 4;4(1):313–23.

28. Olanow CW, Obeso JA, Stocchi F. Continuous dopamine-receptor treatment of Parkinson’s disease: scientific rationale and clinical implications. Lancet Neurol. 2006 Aug;5(8):677–87.

29. Cilia R, Akpalu A, Sarfo FS, et al. The modern pre-levodopa era of Parkinson’s disease: insights into motor complications from sub-Saharan Africa. Brain. 2014 Oct;137(Pt 10):2731–42.

30. Gibson JS, Flanigan JL, Patrie JT, Dalrymple WA, Harrison MB. Predictors of anxiety in Parkinson’s disease: results from a 3-year longitudinal cohort study. Neurol Sci. 2023 Feb;44(2):547–56.

31. Ratajska AM, Etheridge CB, Lopez FV, et al. The Relationship Between Autonomic Dysfunction and Mood Symptoms in De Novo Parkinson’s Disease Patients Over Time. J Geriatr Psychiatry Neurol. 2024 May;37(3):242–52.

32. Blase K, Vermetten E, Lehrer P, Gevirtz R. Neurophysiological Approach by Self-Control of Your Stress-Related Autonomic Nervous System with Depression, Stress and Anxiety Patients. Int J Environ Res Public Health. 2021 Mar 24;18(7).

33. Beauchaine TP, Thayer JF. Heart rate variability as a transdiagnostic biomarker of psychopathology. Int J Psychophysiol. 2015 Nov;98(2 Pt 2):338-50.

34. Alvares GA, Quintana DS, Hickie IB, Guastella AJ. Autonomic nervous system dysfunction in psychiatric disorders and the impact of psychotropic medications: a systematic review and meta-analysis. J Psychiatry Neurosci. 2016 Mar;41(2):89–104.

35. Carli G, Kanel P, Michalakis F, et al. Cardiac sympathetic denervation and anxiety in Parkinson disease. Parkinsonism Relat Disord. 2024 May 3;124:106997.

36. Berrios GE, Campbell C, Politynska BE. Autonomic failure, depression and anxiety in Parkinson’s disease. Br J Psychiatry. 1995 Jun;166(6):789–92.

37. McDonald C, Newton JL, Burn DJ. Orthostatic hypotension and cognitive impairment in Parkinson’s disease: Causation or association? Movement disorders : official journal of the Movement Disorder Society. 2016 Jul;31(7):937–46.

38. Udow SJ, Robertson AD, MacIntosh BJ, et al. ’Under pressure’: is there a link between orthostatic hypotension and cognitive impairment in α-synucleinopathies? J Neurol Neurosurg Psychiatry. 2016 Dec;87(12):1311–21.

39. Ruiz Barrio I, Miki Y, Jaunmuktane ZT, Warner T, De Pablo-Fernandez E. Association Between Orthostatic Hypotension and Dementia in Patients With Parkinson Disease and Multiple System Atrophy. Neurology. 2023 Mar 7;100(10):e998–e1008.

40. Choi S, Kim R, Kang N, Byun K, Park K, Jun JS. Associations of Orthostatic Hypotension and Orthostatic Intolerance with Domain-Specific Cognitive Decline in Patients with Early Parkinson Disease: An 8-Year Follow-up. J Am Med Dir Assoc. 2023 Nov 4.

41. Sara SJ. The locus coeruleus and noradrenergic modulation of cognition. Nat Rev Neurosci. 2009 Mar;10(3):211–23.

42. Espay AJ, LeWitt PA, Kaufmann H. Norepinephrine deficiency in Parkinson’s disease: the case for noradrenergic enhancement. Movement disorders : official journal of the Movement Disorder Society. 2014 Dec;29(14):1710–9.

43. Riley DE, Espay AJ. Cognitive fluctuations in Parkinson’s disease dementia: blood pressure lability as an underlying mechanism. J Clin Mov Disord. 2018;5:1.

44. Centi J, Freeman R, Gibbons CH, Neargarder S, Canova AO, Cronin-Golomb A. Effects of orthostatic hypotension on cognition in Parkinson disease. Neurology. 2017 Jan 3;88(1):17–24.

45. Espay AJ, Marsili L, Mahajan A, et al. Rivastigmine in Parkinson’s Disease Dementia with Orthostatic Hypotension. Ann Neurol. 2021 Jan;89(1):91–8.

46. Seemiller J, Morrow C, Hinkle JT, et al. Impact of Acute Dopamine Replacement on Cognitive Function in Parkinson’s Disease. Mov Disord Clin Pract. 2024 May;11(5):534–42.

47. Pascual-Sedano B, Kulisevsky J, Barbanoj M, et al. Levodopa and executive performance in Parkinson’s disease: a randomized study. J Int Neuropsychol Soc. 2008 Sep;14(5):832–41.

48. Ruitenberg MFL, Abrahamse EL, Santens P, Notebaert W. The effect of dopaminergic medication on conflict adaptation in Parkinson’s disease. J Neuropsychol. 2019 Mar;13(1):121–35.

49. Cools R, Miyakawa A, Sheridan M, D’Esposito M. Enhanced frontal function in Parkinson’s disease. Brain. 2010 Jan;133(Pt 1):225–33.

50. Defer GL, Widner H, Marié RM, Rémy P, Levivier M. Core assessment program for surgical interventional therapies in Parkinson’s disease (CAPSIT-PD). Movement disorders : official journal of the Movement Disorder Society. 1999 Jul;14(4):572–84.

