## Supplementary data for "The effect of dysautonomia on motor, behavioral and cognitive fluctuations in Parkinson’s disease"

**Supplementary Material**

**Supplementary table 1. Linear regression model using the total SCOPA-AUT score (exposure variable) and change scores as continuous variable.**

|  | Covariates | Coefficient (SE) | p-value |
| --- | --- | --- | --- |
| MDS- UPDRS III difference (post vs. pre-treatment) | Dysautonomia | -0.15 (0.08) | 0.06 |
|  | Duration of disease | -0.19 (0.10) | 0.06 |
|  | LEDD | -0.003 (0.001) | 0.03 |
|  | Agonist LEDD | -0.0002 (0.004) | 0.96 |
| SDMT difference (post vs. pre-treatment) | Dysautonomia | 0.14 (0.07) | 0.03 |
|  | Duration of disease | -0.23 (0.09) | 0.01 |
|  | LEDD | -0.0005 (0.001) | 0.64 |
|  | Agonist LEDD | -0.0004 (0.003) | 0.91 |
| STROOP difference (post vs. pre-treatment) | Dysautonomia | -0.17 (0.07) | 0.03 |
|  | Duration of disease | -0.09 (0.10) | 0.38 |
|  | LEDD | 0.0008 (0.001) | 0.49 |
|  | Agonist LEDD | -0.004 (0.004) | 0.34 |
| HAM-A difference (post vs. pre-treatment) | Dysautonomia | -0.13 (0.05) | 0.007 |
|  | Duration of disease | -0.07 (0.06) | 0.25 |
|  | LEDD | -0.002 (0.0007) | 0.03 |
|  | Agonist LEDD | 0.001 (0.002) | 0.56 |
| HAM-D difference (post vs. pre-treatment) | Dysautonomia | -0.02 (0.03) | 0.40 |
|  | Duration of disease | -0.10 (0.03) | 0.004 |
|  | LEDD | -0.0004 (0.0004) | 0.33 |
|  | Agonist LEDD | -0.001 (0.001) | 0.44 |

**Supplementary Figure:** Plot of Logistic Regression Model Assessing Impact of dysautonomia on statistically significant, clinically-important Fluctuations in Parkinson’s disease

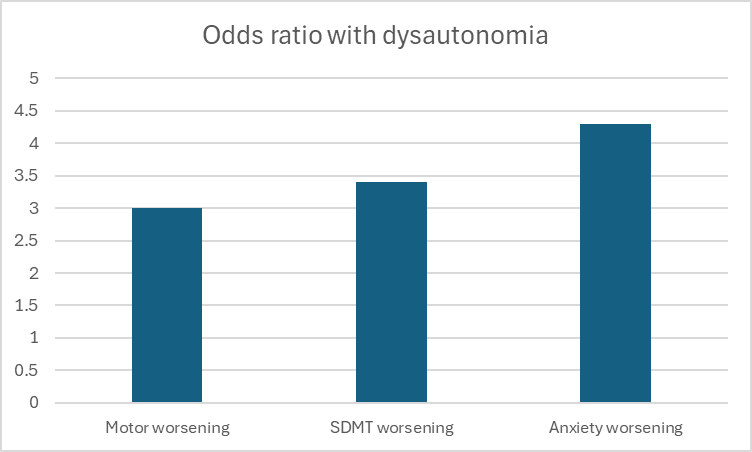

**Supplementary table 2.** Dysautonomia on medication-induced changes in motor and non-motor features: raw data on change in scores

|  | DYS-  (n = 94) | DYS+  (n = 106) | t-value | *p*-value |
| --- | --- | --- | --- | --- |
| Cognition |  |  |  |  |
| Stroop test score Off | 34.2 (9.3) | 32.7 (12.9) | 0.87 | 0.38 |
| Stroop test score On | 37.1 (11.3) | 33.8 (12.9) | 1.8 | 0.08 |
| Stroop difference (On – Off) | 2.7 (6.9) | 1.0 (6.2) | 1.7 | 0.09 |
| SDMT score Off | 48.0 (11.0) | 41.7 (12.9) | 3.5 | <0.001 |
| SDMT score On | 46.0 (11.0) | 40.7 (14.0) | 2.8 | 0.006 |
| SDMT difference (On – Off) | -2.3 (5.9) | -0.91 (6.6) | -1.5 | 0.13 |
| Motor Symptoms |  |  |  |  |
| MDS UPDRS III score Off | 37.3 (13.6) | 42.8 (15.1) | -2.7 | 0.008 |
| MDS UPDRS III score On | 25.1 (11.3) | 28.4 (13.1) | -1.9 | 0.06 |
| MDS UPDRS III difference (On – Off) | -12.2 (8.1) | -14.3 (7.6) | 1.9 | 0.05 |
| Psychiatric Symptoms |  |  |  |  |
| HAM-A score Off | 10.2 (5.9) | 17.1 (15.7) | -7.2 | <0.001 |
| HAM-A score On | 6.2 (4.3) | 11.1 (5.2) | -7.2 | <0.001 |
| HAM-A difference (On – Off) | -4.0 (4.7) | -6.0 (4.8) | 2.9 | 0.004 |
| HAM-D score Off | 5.0 (4.2) | 8.6 (5.9) | -4.9 | <0.001 |
| HAM-D score On | 3.4 (3.1) | 6.2 (5.2) | -4.5 | <0.001 |
| HAM-D difference (On – Off) | -1.5 (2.6) | -2.3 (2.6) | 2.1 | 0.04 |
